# Increased aperiodic offset and heightened alpha power characterize resting-state EEG activity in Parkinson’s disease

**DOI:** 10.64898/2026.01.20.26343832

**Authors:** Salvatore Bertino, Amin Ghaderi-Kangavari, David Meder, Mikkel Christoffer Vinding, Nora Raaf, Lasse Christiansen, Birgitte Liang Chen Thomsen, Annemette Løkkegaard, Angelo Quartarone, Mikkel Malling Beck, Hartwig Roman Siebner

## Abstract

Parkinson’s disease (PD) is characterized by widespread neurodegeneration across neuromodulatory systems, profoundly affecting cortico–basal ganglia–thalamic loops. The resulting cortical alterations can be detected with electroencephalography (EEG). Emerging evidence shows that non-oscillatory, aperiodic EEG activity provides meaningful information that complements the oscillatory, periodic features traditionally examined. Nonetheless, available information on periodic alpha and beta power as well as aperiodic features in PD patients during the OFF-medication state are inconclusive. Moreover, replicability of spectral EEG findings across different EEG systems has received little attention. Herein, we aimed at characterizing aperiodic and periodic EEG signatures in PD during OFF-medication resting-state and examining their relationships with age, disease duration, and levodopa-induced dyskinesias. Resting-state EEG was recorded from forty patients with PD OFF-medication and twenty-six age-and sex-matched healthy controls, and a subset of patients and controls returned for a second visit using a different EEG amplifier. Power spectra were parametrized to extract aperiodic parameters (exponent and offset, i.e. the slope and the intercept of the broadband signal, respectively) and power in the alpha and beta frequency bands, adjusted for the aperiodic component. Bayesian statistical tests were used to assess between-group differences and associations with clinical (MDS-UPRDS-III, LEDD, disease duration) and demographic (age) variables. We found that aperiodic offset and alpha power were higher in patients than controls across all channels (BF_10_=80.74; BF_10_=26.81) and when restricting analysis to pericentral channels (BF_10_=70.75; BF_10_=16.90). These differences were equally expressed in patients with and without a positive history of levodopa-induced dyskinesias and replicated at follow-up using a different EEG recording system. In patients, aperiodic offset and exponent, but not periodic alpha or beta power, scaled positively with age and disease duration (BF_10_=2.16–9.76). The successful replication of increased aperiodic offset and periodic alpha power highlights their potential as robust neurophysiological markers of PD, while aperiodic parameters may track disease duration.

## Introduction

Parkinson’s disease (PD) is a neurodegenerative disorder causing motor and non-motor symptoms, arising from complex interactions of neurodegeneration(1), medication(2), and compensatory mechanisms(3,4) affecting large-scale subcortical-cortical circuits(5,6). Alterations in cortico-basal ganglia thalamocortical loops and neurotransmitter systems(6,7) influence cortical activity, which can be captured using electroencephalography (EEG) and may feasibly provide potential disease markers, especially in the OFF-state, i.e., when cortical activity is not directly or indirectly influenced by dopaminergic medications.

EEG or MEG studies performing traditional spectral analysis on resting-state data in PD mostly report an increase in narrow-band power in the delta-theta range (8–10). Power changes of resting-state activity in the alpha and beta bands are inconsistent across studies: While some studies report no differences in the alpha band between healthy individuals and non-demented PD patients (9,11), others report either increase(12,13) or decrease(14,15) in alpha power. Likewise, the available literature reports either increased (13), decreased (11) or no differences (15) in cortical beta band power between patients and controls. Moreover, such narrow-band alterations have been mostly linked to cognitive disturbances and other non-dopaminergic symptoms(16–19), whereas associations with cardinal motor symptoms of PD (e.g. bradykinesia, rigidity, tremor) remain inconsistent (17,20).

From a methodological standpoint, a key limitation of conventional spectral analyses is that they often overlook the EEG aperiodic component, which contributes broadband power and can confound interpretations of oscillatory changes(21). Hence, alterations in aperiodic activity may underlie apparent differences in periodic power, including the above-described narrow-band changes (22).

Recently introduced spectral parametrization method conceptualizes power spectra as a combination of periodic (i.e oscillatory) and aperiodic components; Such technique leverages on an algorithm that fits models to power spectra to separately and explicitly measure their aperiodic (i.e., *exponent* and *offset*) and periodic (i.e., *peak frequency*, *bandwidth*, and *power*) features (23,24) allowing a more detailed characterization of cortical activity changes in patients populations(25). Such method is agnostic to the way power spectra have been obtained but relies on a priori assumptions on the number of spectral peaks, minimum peak height and width, presence/absence of a knee in the aperiodic component(23). Thus, interpretation of its parameters strongly depends on patient features (e.g. cognitive status, medication state, brain state), data modality (EEG, MEG, local field potentials obtained from intracranial EEG or deep brain stimulation) as well as methodological factors such as EEG system, preprocessing of EEG data, power spectra computation. All these variables encourage testing the replicability of results and their robustness to methodological factors.

Recent resting-state EEG studies jointly examining periodic and aperiodic components in PD, reported divergent results (26–28). Specifically, Wang and colleagues described increased theta power among individuals affected by PD but no differences in aperiodic parameters between patients during their OFF-state and healthy controls (27). Conversely, McKeown and colleagues found that aperiodic exponent, offset and periodic alpha power during eyes open were significantly increased in individuals affected by PD during their OFF-state (26). Finally, Monchy and colleagues restricted their analysis to aperiodic parameters, describing increased aperiodic offset but not aperiodic exponent among PD patients during while being on their regular therapeutic regimen (28).

Current evidence on EEG spectral alterations in PD, including studies applying spectral parameterization approaches, is primarily derived from relatively small samples characterized by heterogeneous medication states and cognitive profiles. In addition, prior work has generally been restricted to a single recording session or EEG system, with little systematic evaluation of the replicability of spectral parameter estimates or of the impact of methodological variability on their measurement. Moreover, the potential influence of a history of levodopa-induced dyskinesias (LID) on periodic and aperiodic EEG activity in the OFF-medication state has not yet been investigated.

To clarify whether alteration of aperiodic and periodic EEG parameters reflect disease-related cortical alterations, we parametrized resting-state EEG power spectra from patients with PD after overnight levodopa withdrawal and age-and sex-matched healthy controls (HC). We aimed to test whether a positive history of peak-dose LID was associated with changes in spectral parameters during the OFF-medication state. We quantified both aperiodic (exponent and offset) and periodic components (alpha and beta power relative to aperiodic component) and analyzed them by averaging across channels. To further investigate potential associations between changes in spectral features and motor symptoms severity, we extracted spectral parameters of pericentral, sensorimotor electrodes as such regions showed altered spectral parameters in recent resting M/EEG studies on PD patients (27,29) while being implicated in expression of cardinal motor symptoms of PD as well as potentially capturing abnormalities peculiar of peak-dose LID (30–32). Hence, parameters obtained from pericentral electrodes were also related to clinical and demographic variables. Finally, spectral parametrization was carried out, using the same pipeline analysis on resting-state EEG data obtained from a subset of subjects that returned for a second session, and using a different EEG acquisition system, to explore robustness of the results and their susceptibility to methodological variables.

## 2. Materials and methods

Forty individuals with PD and twenty-six age-matched healthy controls participated in the study. Patients had no history of neurologic and psychiatric diseases other than PD and were not on medications commonly considered to act on the central nervous system except dopaminergic treatment. Twenty PD patients had a positive history of peak-dose LID derived from previous neurological examinations. The study was carried out in accordance with the Declaration of Helsinki and approved by the local ethics committee of the Capital Region of Denmark, as part of a larger study aimed at investigating neurophysiological and neuroimaging underpinnings of bradykinesia and dyskinesia in PD (H-22010296). All participants provided written informed consent before participating in the study.

Patients were studied in a pragmatic OFF-medication state after overnight (>12 hours) withdrawal from dopaminergic medications. A comprehensive clinical assessment was conducted at enrollment, including the Montreal Cognitive Assessment (MoCA)(33) and the non-motor symptoms scale (NMSS)(34). Dopaminergic doses were converted to levodopa equivalent daily dose (LEDD)(35). Before the EEG measurements, patients were clinically evaluated using the Movement Disorders Society-Unified Parkinson’s Disease Rating Scale subscale III (MDS-UPDRS III)(36). Clinical and demographic data are summarized in Table 1.

**Table 1.** Demographics and clinical scores. Patients with PD were divided into two groups depending on the presence of levodopa-induced dyskinesias (LID). Data are reported as mean (standard deviation). BF_10_ resulting from each statistical analysis are reported in the right-most column.

|  | Healthy Controls | PD patients without LID | PD patients with LID | BF |
| --- | --- | --- | --- | --- |
| Number of subjects | 26 | 20 | 20 |  |
| Age (years) | 64.5 (9.5) | 67.1 (8.9) | 60.65 (8.9) | 0.79 |
| Sex (female/male) | 13/13 | 10/10 | 11/9 | 0.12 |
| Disease Duration (years) | N/A | 6.3 (2.9) | 7.6 (4.6) | 0.48 |
| MoCA score | 27.7 (1.7) | 27.4 (2.4) | 28 (1.8) | 0.16 |
| MDS-UPDRS-III (OFF) | N/A | 30.3 (10.7) | 37.3 (11.0) | 2.07 |
| Hoehn&Yahr | N/A | 2.1 (0.55) | 2.3 (0.41) | 0.44 |
| LEDD | N/A | 639 (270) | 853 (484) | 0.97 |
| NMSS score | N/A | 41.6 (24.5) | 51.6 (27.7) | 0.54 |

### 2.2 EEG recording and pre-processing

Each participant underwent a three-minute resting-state EEG recording with eyes open using 128 channels linked to a BioSemi ActiveTwo EEG system (BioSemi, Amsterdam, Netherlands). The duration of EEG recordings, comparable to available studies in the field (26–28), was motivated by the need of preventing spectral slowing due to sleepiness while ensuring at least two minutes of artifact-free data after data-preprocessing. Data were collected with a sampling frequency of 2048 Hz. During the recordings, participants were asked to relax and fixate their gaze at a cross on a monitor approximately one meter in front of them. A subset of subjects (25 patients, 13 of them with a positive history of LID and 10 healthy controls) underwent a second EEG recording, on a separate day, where data were acquired from 61 passive Ag/AgCl electrodes placed in an equidistant EEG cap (M10 cap layout, BrainCap TMS, Brain Products GmbH, Germany), collected at a sampling frequency of 50000 Hz (actiCHamp Plus 64 System, Brain Products GmbH, Germany). In both cases, during cap preparation, impendences were held equal or inferior to 5 kOhm for each electrode.

The same preprocessing pipeline was implemented in PD patients and healthy controls using the MNE-Python toolbox (https://mne.tools)(37). EEG signals were first downsampled to 1000 Hz and bandpass filtered between 0.1 and 45 Hz. All channels were visually inspected, and those characterized by high-amplitude artifacts consistent across trials were removed and spherically interpolated. After interpolation, data were re-referenced to the average reference and segments containing noise (e.g., muscle activity affecting all the channels) were visually identified and excluded.

Independent component analysis (ICA) using the FastICA algorithm(38) was applied to identify and remove components of non-brain origin. To enhance spatial specificity at the sensor level, a surface Laplacian filter was applied (stiffness *m=5,* Legendre polynomial = 80, λ = 10^-3^), where *m* represents the smoothness of the results, Legendre polynomial measures the spatial specificity of the results, and λ refers to noise suppression (39,40). These parameters were selected due to the higher noise level typically present in EEG recordings from PD patients during the OFF condition, which can complicate the isolation of physiologically relevant features (41).

### 2.3 Power spectra computation and parametrization

The power spectral density (PSD) for each EEG channel was estimated on preprocessed data using Welch’s method (window length: 2 seconds, frequency resolution 0.5 Hz, window overlap 50%). Power spectra were then parametrized using the Fitting Oscillations and One-Over-F (FOOOF) toolbox(23), which models the power spectral density as a combination of aperiodic and periodic components. The model was initialized with the following parameters: peak width limits = 1-8 Hz, maximum number of peaks = 2, peak threshold = 1, minimum peak height = 0.1, with the aperiodic mode set to “fixed’’, i.e. without accounting for the presence of a bend in the log-log space, as such feature is more frequently employed with larger frequency intervals (e.g. local field potentials LFPs from invasive recordings) (23). We chose our model parameters based on those used in previously published studies(23,42), specifically, we constrained our analysis to maximum two peaks, as resting-state EEG is not expected to contain more than two prominent oscillatory peaks. This approach reduces the risk of overfitting and avoids detecting small overlapping peaks that may not represent genuine oscillations. We evaluated the model performance by visual inspection of power spectra for each subject, along with quantification of goodness-of-fit. Specifically, we computed *R^2^*, a measure of correspondence between the original power spectrum and the model fit and the mean absolute error between the model fit and the original power spectrum(23). We found almost clean-cut visual correspondence between the raw and parameterized spectra, together with high R^2^ averaged across channels and participants, with values nearly identical for healthy controls and PD (mean: 0.97; STD: 0.07).

The aperiodic component is defined by the exponent (slope) and offset (intercept with the y-axis) of the log-transformed power spectrum. Adjusted periodic features were extracted as power above the aperiodic background in canonical frequency bands(43): alpha (7–15 Hz) and beta (15–35 Hz).

For further analyses, (i) spectral parameters were averaged across all channels, and (ii) extracted from two electrodes overlying left and right sensorimotor regions (C3 and C4, according to the international 10-20 EEG system(44)), which were averaged to yield a representative sensorimotor signal (see Figure 1 for a summary of: preprocessing pipeline and spectral parameterization workflow).

**Figure 1.**
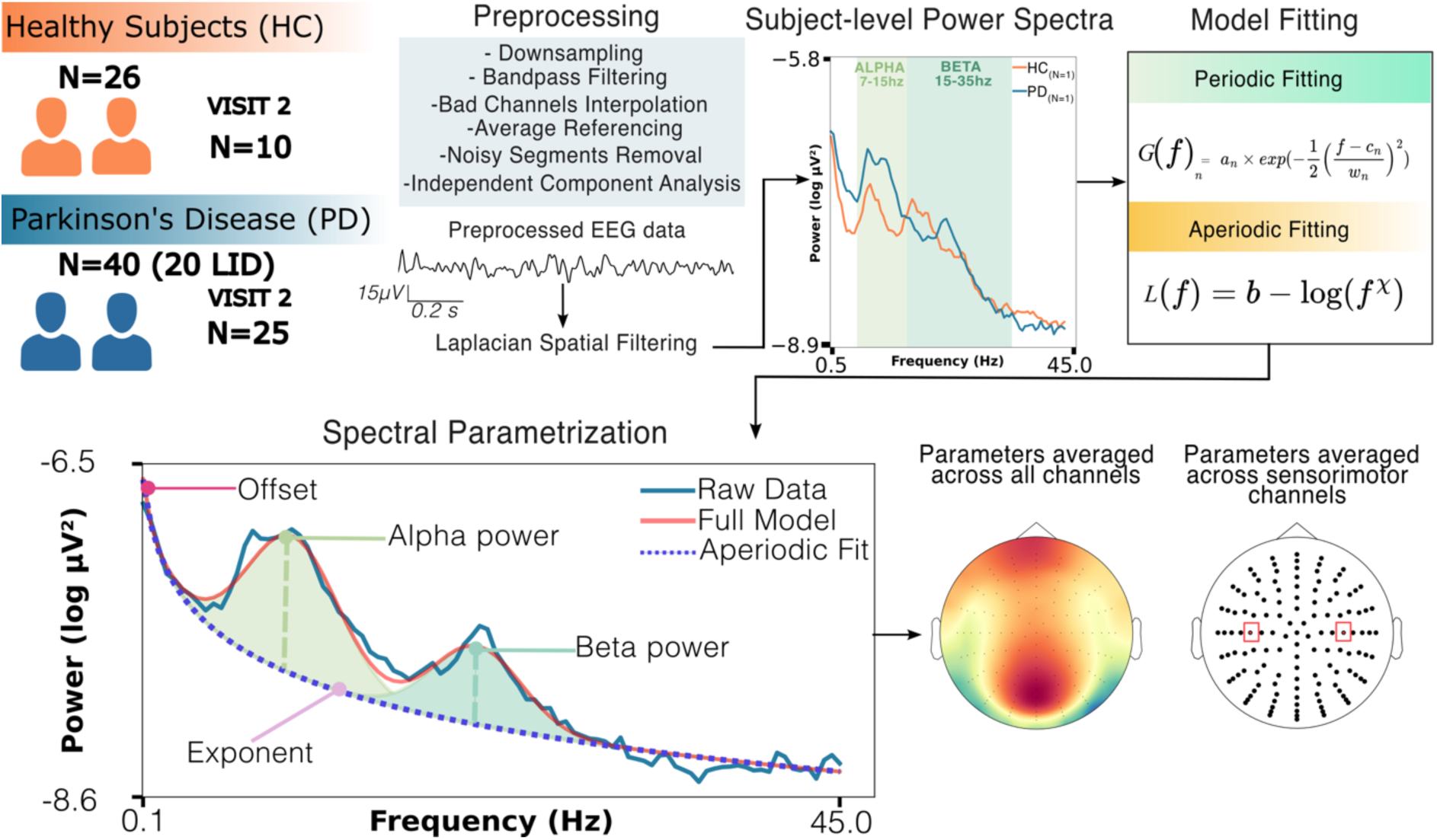
Pre-processing steps of single-subject resting-state EEG data to derive periodic and aperiodic components. Power spectra were computed for each EEG channel using the Welch’s method (exemplary data from channel C3 of one PD patient and one healthy control) and parametrized using the fitting oscillation and one over f (FOOOF) toolbox (top right panel): The periodic component is modelled using Gaussian functions where a is the height of the peak, c represents the centre frequency, w is the peak width and f is frequency. The aperiodic component is modelled using a Lorentzian function where b is the offset, **_χ_** is the exponent, and f represents frequency. Parameterization of power spectra allowed the distinct estimation of the aperiodic component (dark blue dotted line) which has been characterized by measuring the aperiodic exponent (or spectral slope, pink) and offset (the overall vertical displacement, or the intercept of the curve with the y-axis, magenta) as well as alpha (light green, dotted line) and beta (dark green, dotted line) power above the aperiodic component.

### 2.4 Statistical analysis

Between-group differences in demographic features (gender, age) as well as clinical variables (MoCA, UPDRS-III, Hoehn&Yahr, LEDD, NMSS) were investigated by Bayesian statistics. Specifically, for the continuous variables available for each group (healthy controls, PD, LID), Bayesian ANOVA were carried out. Conversely, when dealing with variables specific to patients with or without history of dyskinesias (UPDRS-III, Hoehn&Yahr, LEDD, NMSS), Bayesian independent samples t-test were employed to explore inter-group differences. Differences in categorical variables (gender) were investigated using Bayesian contingency tables (see. Table 1).

The spectral parameters averaged across all channels and for sensorimotor channels were compared across healthy controls and PD groups using Bayesian analysis of covariance (ANCOVA). Individual models were fitted with spectral parameters (exponent, offset, alpha power, beta power) as the dependent variables and group (PD vs. healthy controls) as the independent variable, with age as a continuous covariate.

As an exploratory analysis, we separated patients with a presence or absence of a history of LID to investigate whether spectral parameters (dependent variables) showed changes related to subgroup (with and without LID) considering age, disease duration, MDS-UPDRS-III, and LEDD as covariates. All the analyses were performed in JASP (Version 0.19.3, https://jasp-stats.org/).

Bayesian model comparison was conducted using Bayes Factors that quantify the continuous evidence in favor of the alternative hypothesis over the null hypothesis. The winning model was identified as the one with the highest Bayes Factor compared to competing models. After selecting the winning model, we examined the Bayes Factor for the inclusion of each parameter and interaction. The Bayes Factor of a model against the null hypothesis is denoted as BF_m10_, while the Bayes Factor for the inclusion of a parameter against the exclusion of the parameter among all possible models is denoted as BF₁₀. Interpretation of Bayes Factors was based on the Jeffreys conventions (45).

## 3. Results

### 3.1 Group differences in demographic and clinical variables

Bayesian statistics showed only anecdotal evidence (BF_10_ < 1), supporting the absence of differences between groups for each of the demographic variables taken into account (age, sex, MoCA); The same pattern was observed in variables pertaining exclusively to patients, where an anecdotal evidence in favor of a between-group difference was observed for MDS-UPDRS-III between patients with and without a positive history of LID (Table 1).

### 3.1 Group differences in aperiodic EEG parameters

Figure 2 shows power spectra obtained after averaging power spectral densities across channels as well as power spectral densities of pericentral channels averaged across subjects, before spectral parametrization. In the PD group, power spectra averaged across channels showed a vertical broadband power shift (Figure 2A); the same pattern applies to sensorimotor channels (Figure 2B-C). Figure 3 shows between-group differences in aperiodic features of power spectra after parametrization. Specifically, figure 3A illustrates the scalp topographies of the two aperiodic parameters and their channel-wise distributions averaged across participants.

**Figure 2.**
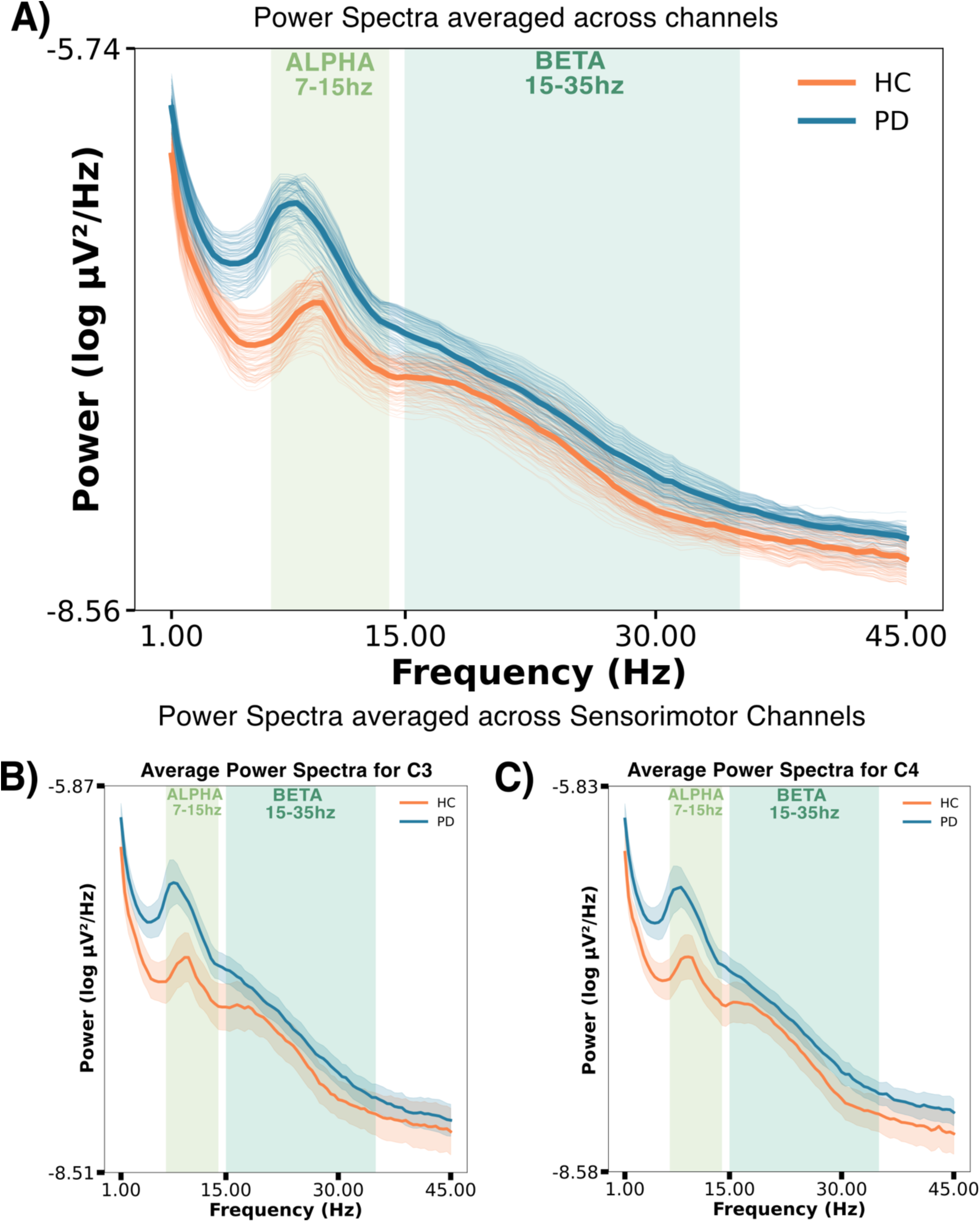
Group-level power spectra derived from power spectral density averaged across all channels and across subjects for sensorimotor channels. The upper panel (A) depicts group-level power spectra obtained log-transforming and averaging power spectral densities across all channels (thick lines) overlaid on average power spectra for each of the 128 channels (thin lines). The lower panel shows group-level power spectra from left (B) and right (C) sensorimotor channels (C3 and C4 according to the 10-20 system). Specifically, power spectral densities of left and right pericentral electrodes overlying sensorimotor areas have been extracted, log-transformed, and averaged across healthy (orange) and individuals with Parkinson’s disease (blue). Group-level, average power spectra are displayed with their respective 95% confidence intervals (orange and blue shaded areas).

**Figure 3.**
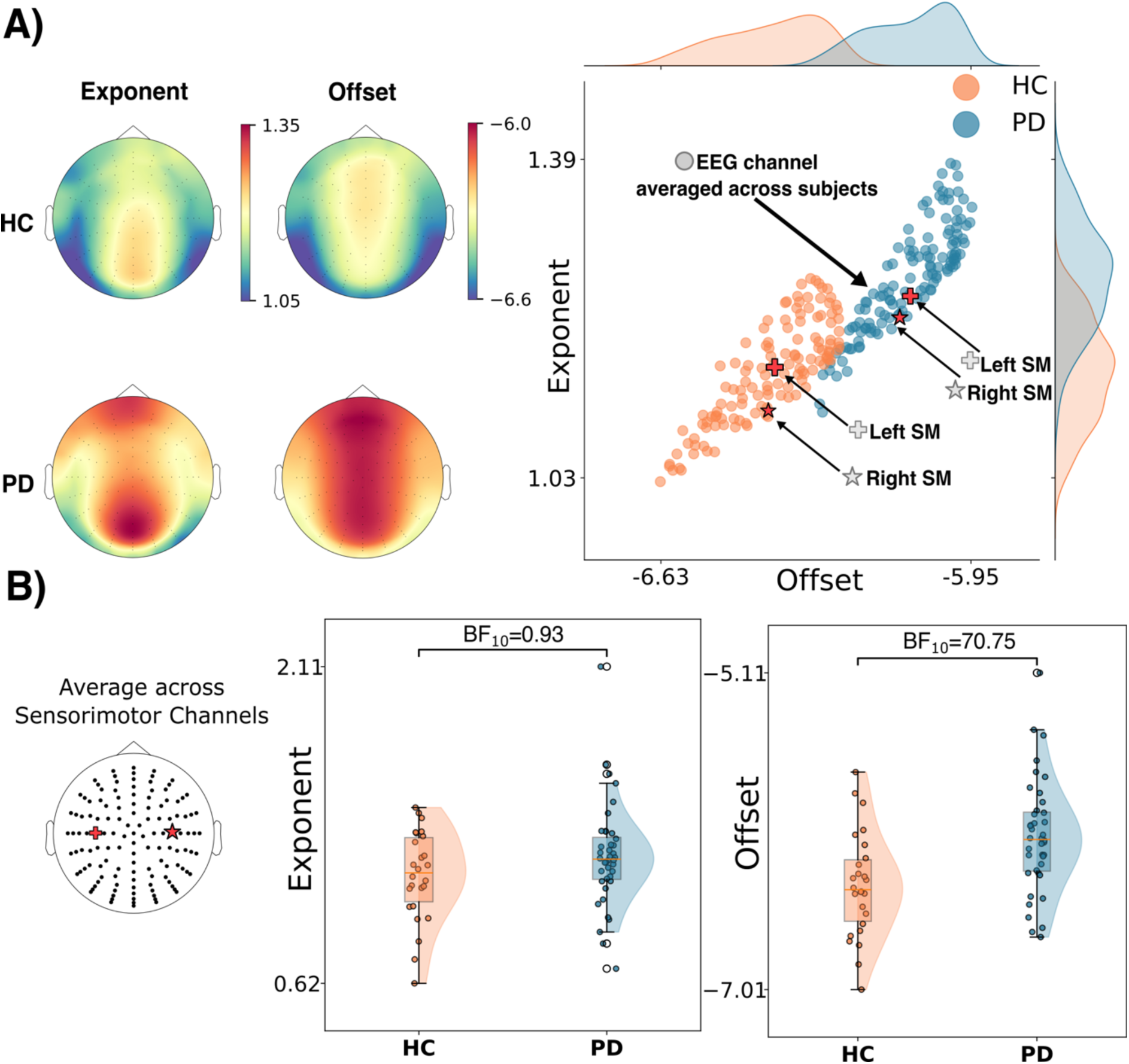
Differences in aperiodic spectral resting-state EEG parameters between PD and healthy controls. (A) Topographic maps show the spatial distribution of exponent and offset for each group at visit 1. Scatterplots depict channel-level grand averages of offset vs. exponent, with marginal density plots showing their distribution across channels. Each scatter represents channel-level exponent and offset averaged across subjects. Left (+) and Right (*) sensorimotor (SM) channels are highlighted and depicted in red as they were selected for further analyses. (B) Raincloud plots of between-group differences in offset and exponent at sensorimotor electrodes. Offsets were higher in PD (group BF₁₀ = 70.75), with no evidence for group differences in exponent (group BF₁₀ = 0.93).

Across all electrodes, there was very strong evidence for higher offset values in the PD group relative to healthy controls; offset also increased with age in both groups (winning model: group+age; BF_m10_= 649.41; group BF_10_=80.74; age BF_10_=18.98, model-averaged posterior coefficient for PD group 0.142, 95% Credible Interval [0.058,0.227]; model-averaged posterior coefficient for control group-0.142, 95% Credible Interval [-0.228,-0.059], model-averaged posterior coefficient for age 0.013, 95% Credible Interval [0.004, 0.021]). In contrast, evidence for group differences in the exponent was anecdotal (group BF_10_=1.61), although exponent values increased with age in both groups (winning model: group+age; BF_m10_=120.31, age BF_10_=96.86; model-averaged posterior coefficient for PD group 0.049, 95% Credible Interval [-0.002,0.102]; model-averaged posterior coefficient for control group-0.049, 95% Credible Interval [-0.103,-0.001], model-averaged posterior coefficient for age 0.010, 95% Credible Interval [0.004, 0.016]).

For subsequent analyses, aperiodic spectral parameters were averaged across electrodes overlying left and right sensorimotor regions (Figure 3B). Consistent with global analysis, there was strong evidence for a higher offset in PD patients (winning model: group+age BF_m10_=288.79; group BF_10_=70.75; age BF_10_=7.64, model-averaged posterior coefficient for PD group 0.136, 95% Credible Interval [0.056,0.217]; model-averaged posterior coefficient for control group-0.136, 95% Credible Interval [-0.218,-0.057], model-averaged posterior coefficient for age 0.011, 95% Credible Interval [0.003, 0.019]), whereas exponents did not show evidence supporting a difference between groups (group BF₁₀ = 0.93) (Figure 3B). The exponent increased with age in both groups (winning model: age; BF_m_₁₀ = 11.88, age BF₁₀ = 13.46; model-averaged posterior coefficient for PD group 0.042, 95% Credible Interval [-0.011,0.094]; model-averaged posterior coefficient for control group-0.042, 95% Credible Interval [-0.095, 0.011], model-averaged posterior coefficient for age 0.008, 95% Credible Interval [0.003, 0.014]).

### 3.2 Group differences in periodic EEG parameters

Bayesian ANCOVA showed very strong evidence for higher alpha power values averaged across all channels in the PD group relative to healthy controls (winning model: group; BF_m10_=25.91; group BF_10_=26.81; age BF_10_=0.35, model-averaged posterior coefficient for PD group 0.136, 95% Credible Interval [0.047,0.227]; model-averaged posterior coefficient for control group-0.136, 95% Credible Interval [-0.228,-0.047], model-averaged posterior coefficient for age 0.004, 95% Credible Interval [0.005, 0.013]) and no evidence for differences in beta power between groups (winning model: null; BF_m10_=1.0; group BF_10_=0.35; age BF_10_=0.24, model-averaged posterior coefficient for PD group-0.027, 95% Credible Interval [-0.032,0.086]; model-averaged posterior coefficient for control group-0.027, 95% Credible Interval [-0.087,-0.031], model-averaged posterior coefficient for age 0.0004, 95% Credible Interval [0.006, 0.007]). In the context of periodic parameters, age did not show any effect on either alpha or beta power. Similar results were observed when restricting the analysis to sensorimotor channels (Figure 4A-B): we observed strong evidence supporting higher alpha power in PD patients (winning model: group; BF_m10_=16.58; group BF_10_=16.90; age BF_10_=0.31, model-averaged posterior coefficient for PD group 0.133, 95% Credible Interval [0.041,0.224]; model-averaged posterior coefficient for control group-0.133, 95% Credible Interval [-0.224,-0.042], model-averaged posterior coefficient for age 0.003, 95% Credible Interval [-0.006, 0.012]), while beta power showed moderate evidence against differences between control and PD groups (winning model: null; BF_m10_=1; group BF_10_=0.42; age BF_10_=0.25, model-averaged posterior coefficient for PD group-0.030, 95% Credible Interval [-0.095,0.031]; model-averaged posterior coefficient for control group 0.030, 95% Credible Interval [-0.095,-0.031], model-averaged posterior coefficient for age 0.0004, 95% Credible Interval [-0.006, 0.007]). As for the average across all channels, age did not affect alpha nor beta power.

**Figure 4.**
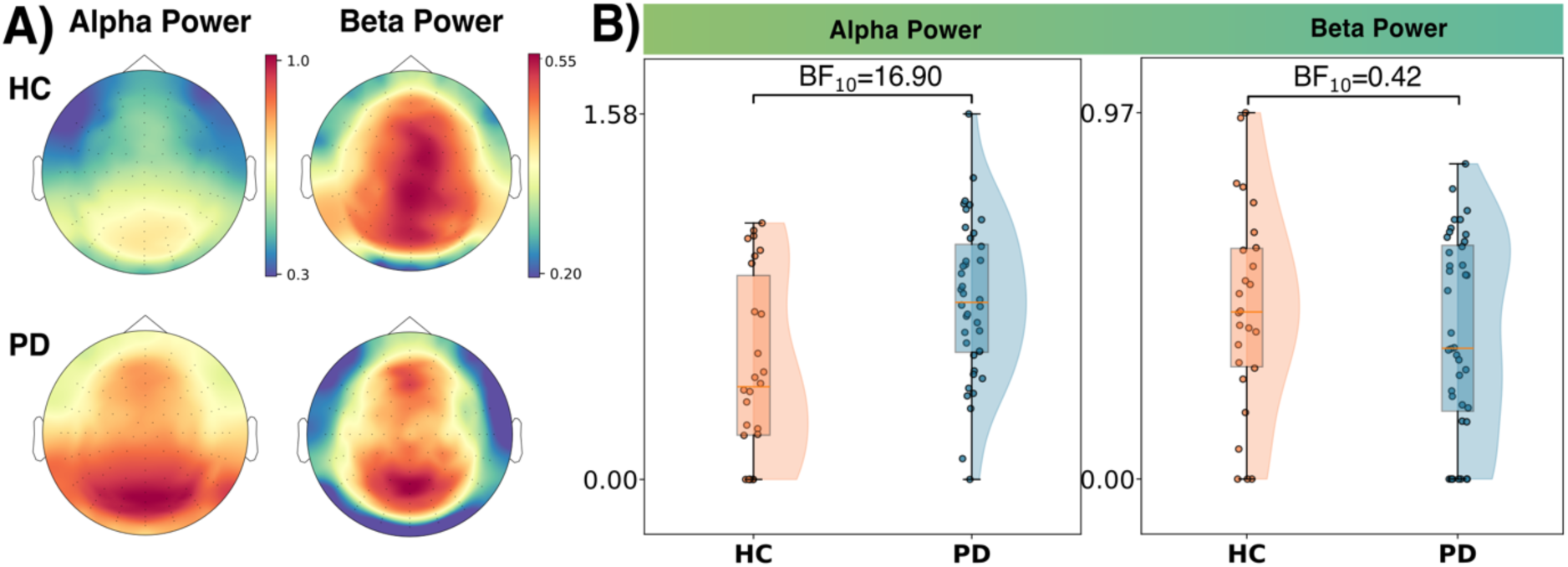
Group differences in periodic EEG parameters within the alpha and beta frequency bands. (A) Topographic maps showing the spatial distribution of alpha and beta power for healthy controls and patients with PD. (B) Raincloud plots show average sensorimotor alpha (left) and beta (right) power for the first session. Bayesian statistics revealed strong evidence for a group difference in alpha power (group BF_10=_16.90) and evidence against a group difference in beta power (group BF_10_=0.42).

As between-group difference in alpha power was topographically widespread, being expressed in the parieto-occipital electrodes but significantly involving also pericentral channels, we sought to investigate whether spectral parameters showed the same pattern when selecting a mid-occipital electrode (Fig.5). Bayesian ANCOVA confirmed the aforementioned pattern, already observed for sensorimotor electrodes as offset was significantly higher in the PD group (winning model: group + age; BF_m10_= 95.21; group BF_10_= 26.07; age BF_10_= 5.87, model-averaged posterior coefficient for PD group 0.139, 95% Credible Interval [0.046,0.230]; model-averaged posterior coefficient for control group-0.139, 95% Credible Interval [-0.231,-0.047], model-averaged posterior coefficient for age 0.012, 95% Credible Interval [0.002, 0.021]), with no meaningful group difference involving the exponent, which increased consensually with age (winning model: age; BF_m10_= 37.41; group BF_10_= 0.96; age BF_10_= 43.27, model-averaged posterior coefficient for PD group 0.051, 95% Credible Interval [-0.012,0.115]; model-averaged posterior coefficient for control group-0.051, 95% Credible Interval [-0.012,-0.115], model-averaged posterior coefficient for age 0.011, 95% Credible Interval [0.004, 0.018]). Neither between-group difference nor covariation with age was observed for beta power (winning model: null; BF_m10_= 1; group BF_10_= 0.32; age BF_10_= 0.31, model-averaged posterior coefficient for PD group:-0.024, 95% Credible Interval [-0.094, 0.045], model-averaged posterior coefficient for control group-0.024, 95% Credible Interval [0.094, 0.045], model-averaged posterior coefficient for age 0.002, 95% Credible Interval [-0.005, 0.010]). Alpha power was higher among PD also in the mid-occipital electrode (winning model: group; BF_m10_= 3.72; group BF_10_= 3.96; age BF_10_= 0.66, model-averaged posterior coefficient for PD group: 0.124, 95% Credible Interval [0.016, 0.231], model-averaged posterior coefficient for control group-0.124, 95% Credible Interval [-0.232,-0.017], model-averaged posterior coefficient for age 0.008, 95% Credible Interval [--0.003, 0.019]) even though the evidence was not as strong as the one observed for alpha averaged across all channels and among left and right sensorimotor channels.

**Figure 5.**
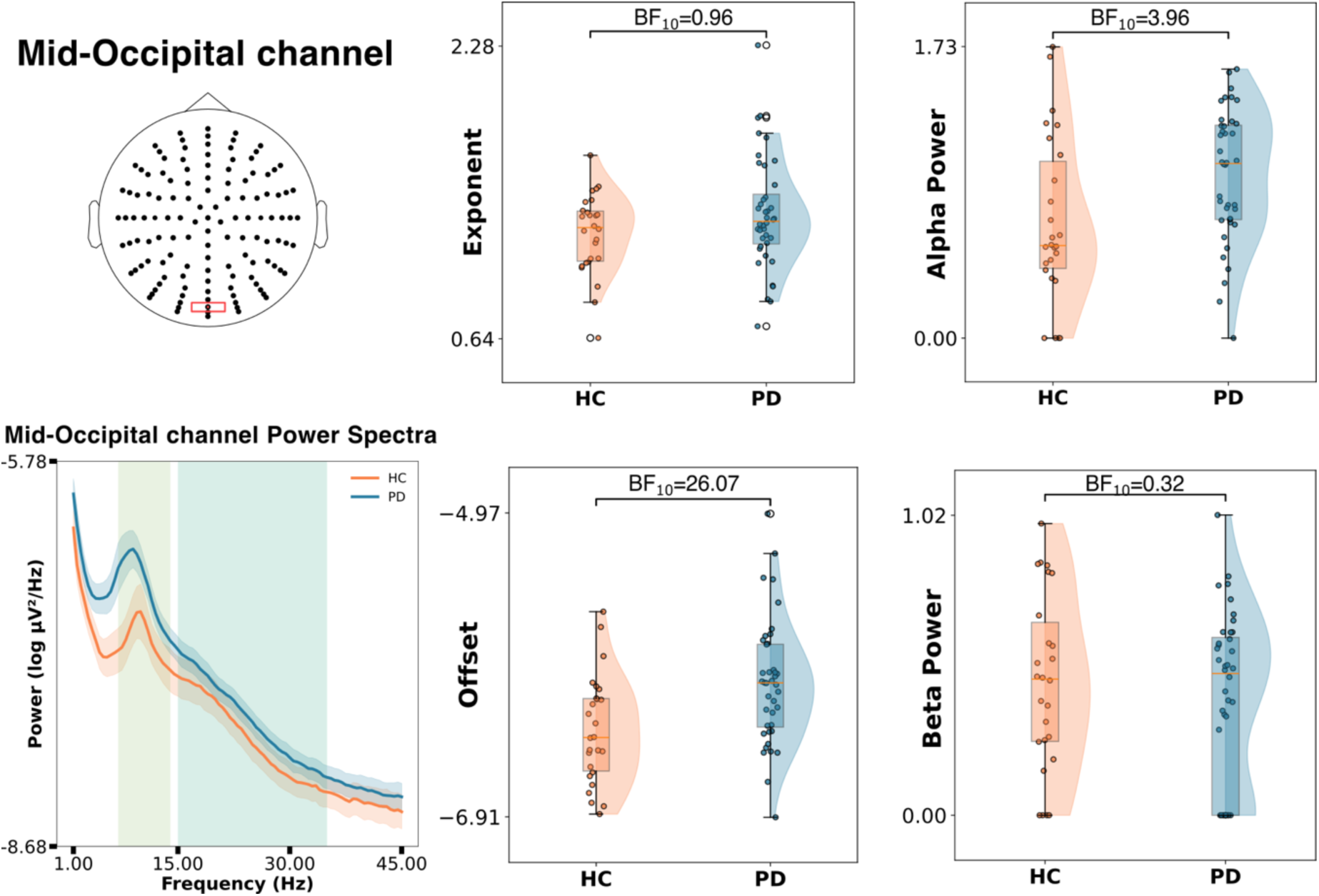
Group differences in aperiodic and periodic EEG parameters at mid-Occipital level. A mid-Occipital electrode corresponding to Oz in the 10-20 system (upper left side of the picture) was selected for further analyses. The lower left side of the picture show power spectra from the mid-occipital channel averaged across subjects for the PD group (blue) and healthy controls (orange), superimposed on their respective confidence interval, prior to parametrization. On the right side of the figure raincloud plots depicting between-group differences in aperiodic and periodic spectral parameters; specifically, as in sensorimotor channels, Offset (group BF_10=_26.90) and alpha power resulted to be higher among patients affected by PD (group BF_10=_3.96). The magnitude of the effect for alpha power is lower than the one observed in sensorimotor channels (see. Fig. 4)

Finally, Given the theoretical orthogonal nature of periodic and aperiodic parameters, we investigated whether the parameters showing significant group differences were correlated. As expected, Bayesian correlation showed no evidence for or against a relationship between offset and alpha power (r = 0.25; BF_10_= 1.065). Notably, these parameters differentiated PD patients from healthy controls, with only little overlap between groups (Figure 6).

**Figure 6.**
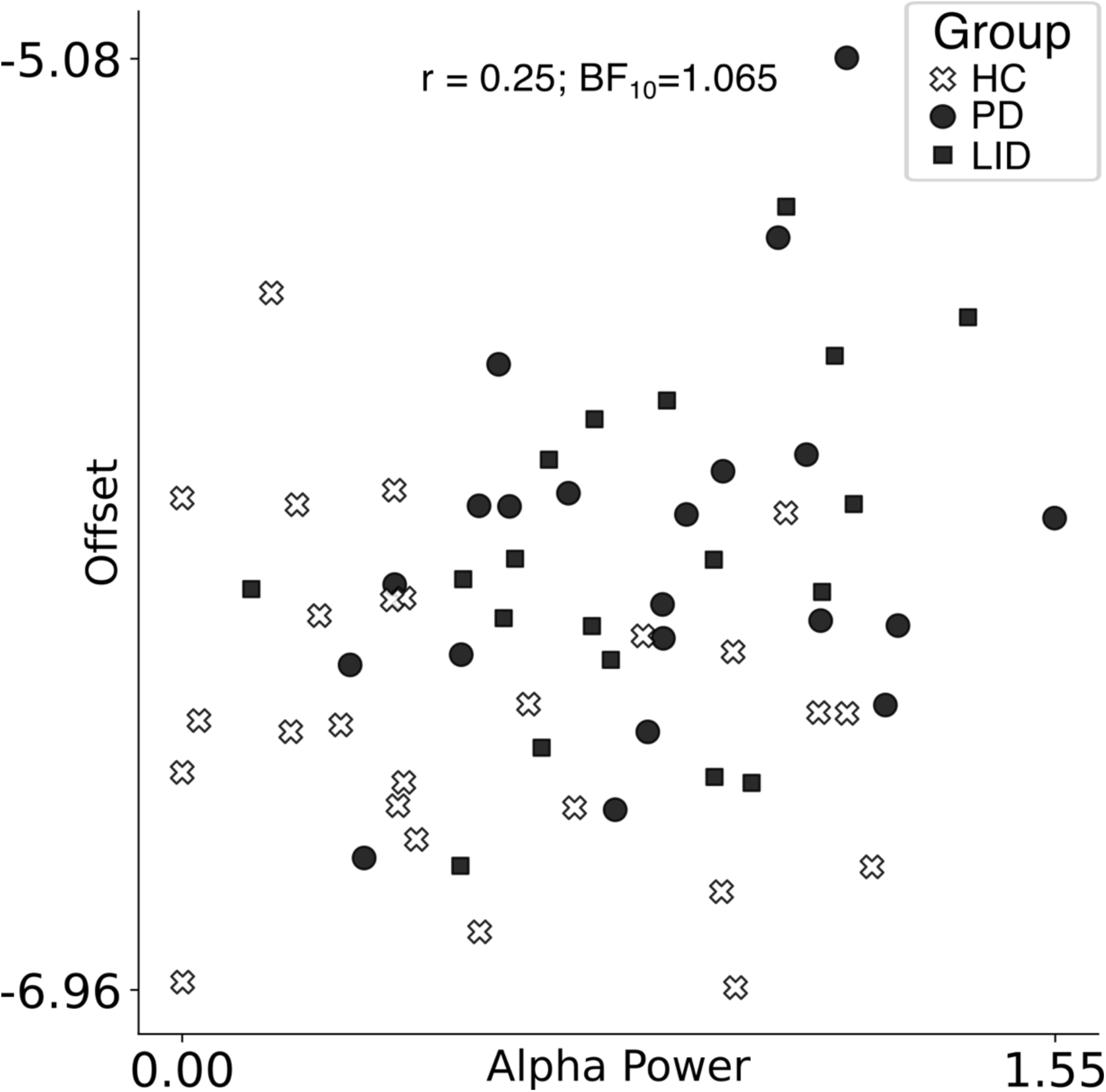
Correlation between aperiodic offset and alpha power. Scatter plot depicting the correlation between aperiodic offset and alpha power, each scatter depicts offset (y-axis) and alpha power (x-axis) for one individual subject, distinguishing between healthy individuals (white crosses) and patients affected by PD either without (black circles) and with a positive history for dyskinesias (black squares). Bayesian correlation showed no evidence in favour or against between alpha power and aperiodic offset (Pearson’s r = 0.25; BF_10_=1.065).

### 3.3 Relationship between spectral EEG parameters and clinical features in individuals with PD

Within the PD group, we compared individuals with (n=20) and without (n=20) a history of LID and examined potential associations with age, disease duration, LEDD and MDS-UPDRS-III. Bayesian ANCOVA provided only low to anecdotal evidence for group differences (PD with LID vs without LID) in both aperiodic and periodic parameters during the OFF-medication state. Specifically, alpha and beta power showed only anecdotal evidence for any effects of covariates, with the null model being favoured in both cases. In contrast, Bayesian ANCOVA revealed effects of age, disease duration, and their interaction on both aperiodic parameters. The winning model (age + disease duration + age × disease duration) was supported for the exponent (BFm₁₀=10.1; age x disease duration BF_10_= 4.09) and offset (BFm₁₀=29.9, age x disease duration BF_10_=5.77). Thus, both aperiodic EEG parameters increased with age and disease duration, with the strongest effects when higher age coincided with longer disease duration (Fig. 7).

**Figure 7.**
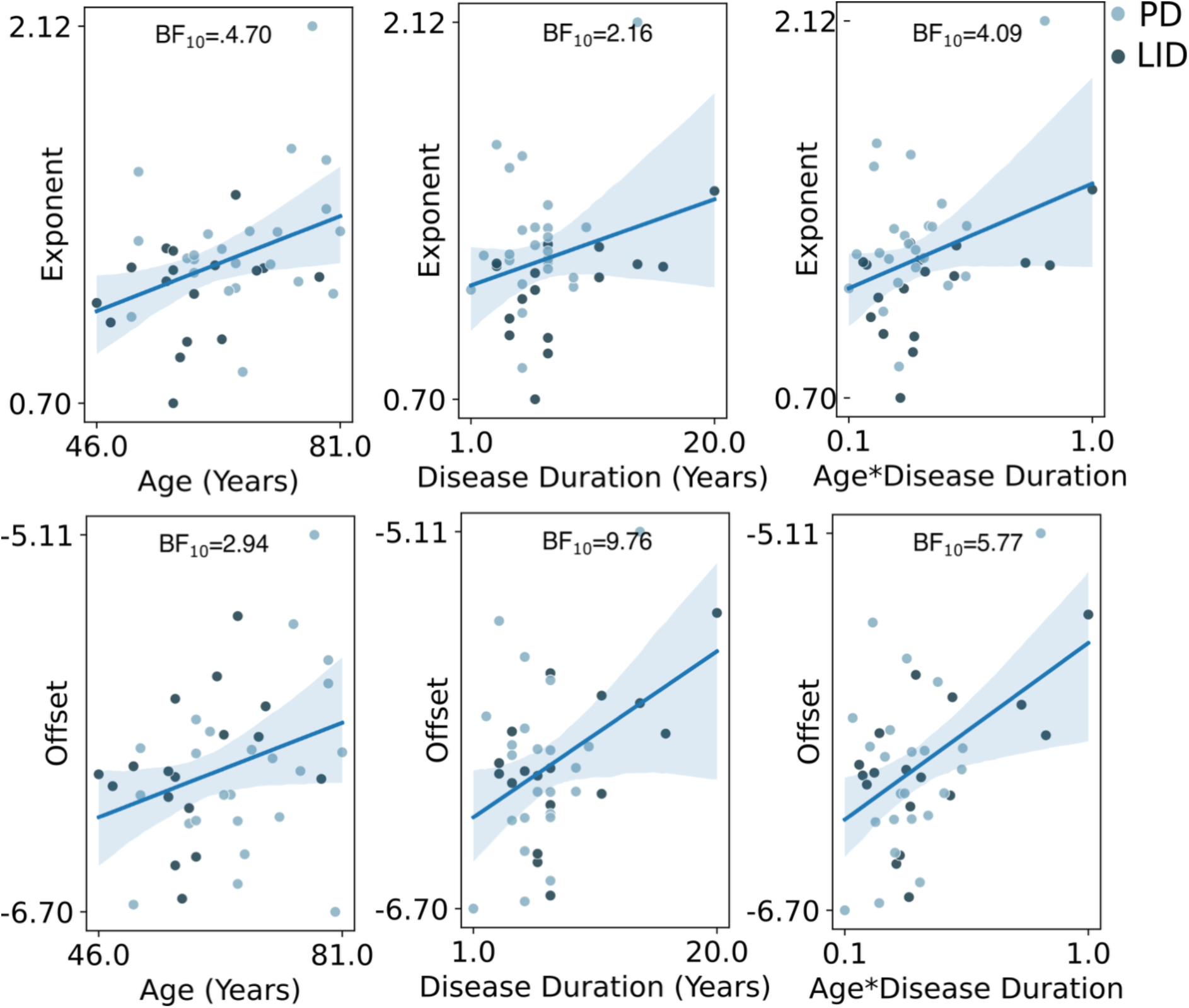
Effects of age and disease duration interact with aperiodic parameters. Scatterplots depicting the effects of age, disease duration and their interaction on both the offset (BF_m10_=29.9) and exponent (BF _m10_=10.1) of the parametrized EEG power spectra of sensorimotor channels. The upper row depicts scatterplots displaying the relation between aperiodic exponent, age (BF_10_=4.70), disease duration (BF_10_=2.16) and their interaction (normalized by dividing the product age*disease duration by the maximum value obtained) (BF_10_=4.1) for patients with (dark blue) and without dyskinesias (light blue). The lower row shows the relation between offset, age (BF_10_=2.94), disease duration (BF_10_=9.76) and their interaction (BF_10_=5.77) for both patient groups.

### 3.4 Group differences replicated on the second visit

Generally, spectral parameters obtained at visit 2 were in accordance the patterns observed during visit 1 (Fig. 8). Notably, aperiodic offset followed a similar topographical distribution to that observed in the first visit (Supplementary Figure 8A). Bayesian ANCOVA, computed over aperiodic features averaged across channels, showed strong evidence supporting a difference between healthy controls and patients for the aperiodic offset (winning model: group+age; BF_m10_=11.25; group BF_10_=7.6; age BF_10_=1.42, model-averaged posterior coefficient for PD group: 0.157, 95% Credible Interval [0.027, 0.292], model-averaged posterior coefficient for control group-0.157, 95% Credible Interval [-0.293,-0.029], model-averaged posterior coefficient for age 0.010, 95% Credible Interval [-0.002, 0.021]) but not for the exponent (winning model: null; BF_m10_=1; group BF_10_=0.86; age BF_10_=0.72, model-averaged posterior coefficient for PD group: 0.064, 95% Credible Interval [-0.034, 0.169], model-averaged posterior coefficient for control group-0.064, 95% Credible Interval [-0.170,-0.033], model-averaged posterior coefficient for age 0.006, 95% Credible Interval [-0.004, 0.016]) replicating results from the first visit. Here, the offset, but not the exponent, scaled with age even though anecdotally. At the sensorimotor level, offset was heightened in PD patients relative to controls (winning model: group; BF_m10_=6.33; group BF_10_=6.12; age BF_10_=0.45, model-averaged posterior coefficient for PD group: 0.157, 95% Credible Interval [0.021, 0.296], model-averaged posterior coefficient for control group-0.157, 95% Credible Interval [-0.297,-0.023], model-averaged posterior coefficient for age 0.004, 95% Credible Interval [-0.007, 0.017]), while the evidence supporting a difference in the exponent between the two groups favoured the null model (winning model: null; BF_m10_=1, group BF_10_=0.74, age BF_10_=0.73, model-averaged posterior coefficient for PD group: 0.065, 95% Credible Interval [-0.045, 0.182], model-averaged posterior coefficient for control group-0.065,, 95% Credible Interval [-0.183, 0.044], model-averaged posterior coefficient for age 0.006, 95% Credible Interval [-0.004, 0.017]) (Figure 8B).

**Figure 8.**
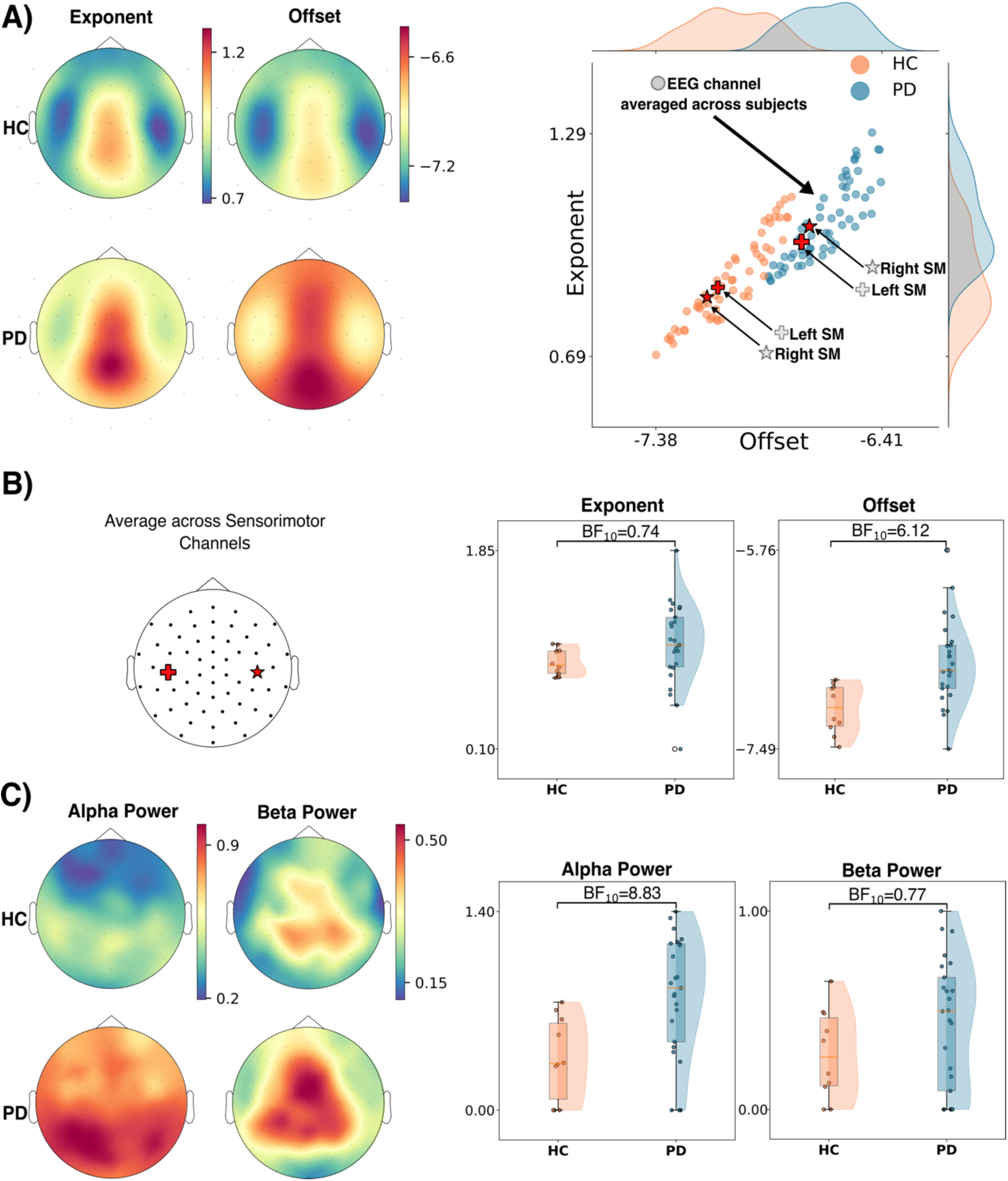
Group differences of aperiodic and periodic resting-state EEG features during visit 2. (A) Panel A displays aperiodic EEG parameters at the sensor level, with topographic maps of exponent and offset across HC and PD patients. Scatterplots display channel-level grand-averages of offset (x-axis) and exponent (y-axis) for each group. (B) The left figure indicates the position of sensorimotor channels in the 61 channels cap. The graph shows raincloud plots showing between-group differences (group BF_10_) when left and right sensorimotor exponent and offset were averaged. (C) This panel depicts topographic maps of alpha and beta power (left panel) and raincloud plots (right panels) summarising group differences in alpha and beta power (group BF_10_) extracted from sensorimotor channels.

Regarding the periodic EEG features, alpha power exhibited a similar parieto-occipital distribution as in Visit 1 (Figure 8C); when averaging across all channels, alpha power showed consistently higher values in the PD group (winning model: group; BF_m10_=58.98; group BF_10_=56.39, age BF_10_=0.39, model averaged posterior coefficient for PD group 0.221, 95% Credible Interval [0.082, 0.356], model averaged posterior coefficient for control group-0.221, 95% Credible Interval [-0.357,-0.083], model averaged coefficient for age 0.004, Credible Interval [-0.008,0.015]), and evidence supporting between-group differences when selecting sensorimotor channels (winning model: group; BF_m10_=8.33; group BF_10_=8.83, age BF_10_=0.562, model averaged posterior coefficient for PD group 0.201, 95% Credible Interval [0.043, 0.361], model averaged posterior coefficient for control group-0.201, 95% Credible Interval [-0.363,-0.045], model averaged coefficient for age 0.006, Credible Interval [-0.007,0.020]). Again, the null-hypothesis was favoured when investigating between-group differences in beta power, both when averaging across channels (winning model: null; BF_m10_=1; group BF_10_=0.64, age BF_10_=0.37, model averaged posterior coefficient for PD group 0.044, 95% Credible Interval [-0.039, 0.134], model averaged posterior coefficient for control group-0.044, 95% Credible Interval [-0.039,0.134], model averaged coefficient for age-0.002, Credible Interval [-0.010,0.006]) and when considering sensorimotor channels (winning model: null; BF_m10_=1; group BF_10_=0.77, age BF_10_=0.35, model averaged posterior coefficient for PD group 0.065, 95% Credible Interval [-0.043, 0.181], model averaged posterior coefficient for control group-0.065, 95% Credible Interval [-0.182,0.042], model averaged coefficient for age 0.002, Credible Interval [-0.009,0.012]) (Figure 8C).

## Discussion

We found that patients with PD exhibited distinct, clinically relevant, periodic and aperiodic features in resting-state EEG compared with age-matched healthy controls. Specifically, patients showed higher aperiodic offset and periodic alpha power, whereas the aperiodic exponent and periodic beta power did not differ between groups. These effects were observed both when averaging across all channels and when extracting parameters from sensorimotor-related pericentral channels. Within the PD group, the increase in aperiodic offset scaled positively with disease duration and age, the latter relationship was also found in healthy individuals. A separate EEG session, carried out on a different day with a different EEG system in a subgroup of participants, replicated these findings, underscoring the robustness of aperiodic offset and alpha power as potential markers of cortical dysfunction in PD.

### Between-group differences in aperiodic offset

The consistently higher spectral offset reconciles previous discrepant findings(26,27), confirming that this effect is evident in the OFF-medication state and likely generalizes across medication conditions. In support of the latter hypothesis, our results also align with a resting-state magnetoencephalography (MEG) study reporting a higher aperiodic offset in PD patients on their regular medication (29). Also, an increased aperiodic offset has also been reported in a recent meta-analysis aggregating EEG/MEG studies employing spectral parametrization on patients affected by PD while being on heterogeneous medication conditions (46). Considered together, these findings suggest that this feature may reflect a stable neurophysiological characteristic of PD rather than a transient dopaminergic effect. Notably, a recent task-based EEG study observed elevated parieto-occipital aperiodic offsets in PD patients during a visuomotor conflict task independent of behavioral performance, showing that aperiodic EEG features and their changes in PD may depend on the ongoing brain state (28). Hence, state-dependent alterations of aperiodic EEG features in PD warrant further studies. From a mechanistic perspective, the neural basis of elevated aperiodic offset likely involves increased broadband neural activity (47). Previous work has linked aperiodic offset to population-level spiking rates, showing that local field potential (LFP) broadband power correlates with neuronal firing rates across cortical regions(48). Although direct evidence in PD is limited, an LFP study on patients with PD, essential tremor and epilepsy reported differences in aperiodic parameters between cortical and subcortical regions, although the offset itself was not the focus(49). A recent multicenter study applying spectral parametrization to LFPs recorded from subthalamic nucleus (STN) of PD patients treated with deep brain stimulation (DBS) found that the aperiodic offset was most strongly associated with clinical symptom severity (50). Moreover, combining the aperiodic offset with oscillatory power in the low-beta (13-18 Hz) and low-gamma (29-45 Hz) frequency ranges explained motor impairment more effectively than either beta power alone or beta power combined with theta and low-gamma power (50). In addition, a measure of broadband spectral power derived from both the aperiodic offset and exponent emerged as a promising within-subject biomarker that was largely independent of medication state (50). These findings suggest that broadband spectral power may reflect increased neuronal spiking activity within the STN of the more affected hemisphere, consistent with observations from animal models of PD (51). Our findings may therefore reflect abnormal spiking activity within the STN that propagates through cortical-basal ganglia circuits, leading to widespread alterations in cortical excitability and/or compensatory neural activity at the cortical level(51–53). However, the neurophysiological mechanisms underlying the observed shifts in cortical spectral offset remain unclear and warrant further investigation. Integrating cellular-and microcircuit-level recordings with in vivo EEG or MEG measurements, together with computational modeling approaches, may help bridge the mechanistic gap between subcortical LFP findings and scalp electrophysiological signatures.

### Relationship between aperiodic EEG parameters and age

We also observed that aperiodic spectral offset scales with age in both PD patients and healthy controls. Age-related changes of aperiodic offset have been poorly investigated in existing studies. One study comparing young and old healthy individuals found no difference in the aperiodic offset(54). Conversely, a study adopting a similar cross-sectional design found lower offsets among older subjects(55). Our findings suggest that age-related increases in aperiodic offset might reflect increased neural activity (52) potentially linked to a para-physiologic compensatory hyperactivity that becomes exaggerated in PD because of abnormalities specifically linked with the disease. Longitudinal studies are needed to clarify whether and how offset changes track healthy aging and its interplay with neurodegeneration.

### No differences between PD patients and controls in the aperiodic exponent

The exponent of the aperiodic power spectrum has been proposed to reflect the excitation-inhibition balance, with steeper slopes indicating greater inhibition(56,57). Unlike the offset, the aperiodic exponent did not differ between groups. This negative finding contrasts with reports of steeper exponents in studies conducted on PD patients (26,27,58,59) and may indicate that an altered excitation–inhibition balance is not a consistent feature of resting EEG in PD. While we cannot exclude the influence of factors such as peculiarities of the clinical population at hand (see(28)) as well as the impact of specific methodological choices (e.g. Laplacian filtering) on exponent, our results were replicated among two different sessions acquired with different EEG systems and align with transcranial magnetic stimulation(60) and magnetic resonance spectroscopy studies(61) showing reduced intracortical inhibition and unchanged GABA levels in PD. Together, these findings argue against enhanced cortical inhibition as a defining electrophysiological feature of the disease.

The winning model from Bayesian ANCOVA suggests that the aperiodic exponent scales with age in our sample, opposing reports of age-related flattening of spectral slopes in healthy individuals (54,55,62,63). It is important to note that studies addressing how aperiodic parameters relate to age were conducted cross-sectionally, comparing groups of young and old healthy individuals, preventing tracking of spectral parameters within subjects. In contrast, the present study focused on a relatively narrow age span and aligns with the findings reported by Helson and colleagues who reported that aperiodic parameters increase as a function of age(58). On the other hand, longitudinal EEG data suggest that the interaction between exponent, alpha peak frequency, and cognitive performance is complex and nonlinear(64), cautioning against simple interpretations of exponent changes as markers of ageing or cognitive decline(64).

### Higher periodic alpha activity in patients with PD

Among patients with PD, we also observed a widespread increase in alpha power in the resting-state EEG, including the pericentral regions. This finding concurs with a recent meta-analysis reporting increased alpha power together with a slowing of alpha peak frequency in PD (46). Given the broad topographical distribution of these effects, we extended our analysis to a mid-occipital electrode, representing a scalp region where the classical posterior alpha rhythm is most prominently expressed (65). At the mid-occipital site, both periodic and aperiodic parameters exhibited patterns like those observed over sensorimotor channels; However, the magnitude of the group difference in alpha power was less prominent than over sensorimotor regions.

One possible interpretation is that the observed alpha-band alterations primarily involve the alpha component of the sensorimotor mu rhythm rather than the classical posterior alpha rhythm. Indeed, impaired mu-rhythm desynchronization has previously been reported in PD(66). This interpretation should, however, be considered with caution. The mu rhythm is characterized by a complex, non-sinusoidal waveform, and its specific properties cannot be adequately resolved using the spectral analyses employed in the present study. Accordingly, future studies need to apply methodological approaches that are specifically designed to characterize sensorimotor mu-rhythm to determine whether alterations in this rhythm contribute to the increased alpha power commonly observed in conventional spectral analyses of PD.

Interestingly, a recent MEG study of patients with PD linked elevated alpha power in the precentral and postcentral gyri of PD patients to locus coeruleus degeneration and attentional deficits, suggesting a contribution of noradrenergic dysfunction to the increased expression of alpha power in the sensorimotor cortex(67). This interpretation aligns with postmortem evidence showing degeneration of brainstem nuclei, affecting neuromodulatory neurotransmitter systems in PD (68,69). Together, these findings raise the possibility that the increased alpha activity observed in our cohort reflects not only alterations in sensorimotor network dynamics but also broader disruptions of neuromodulatory systems.

### No differences between PD patients and controls in the periodic beta EEG component

In contrast, resting-state EEG recordings yielded no evidence for differences in beta power between patients with PD and healthy controls. This negative finding is consistent with with a growing body of literature suggesting that static measures of beta power have limited sensitivity for discriminating disease status or clinical severity in PD (29,70–72). Instead, increasing evidence indicates that the temporal dynamics of beta activity, particularly beta-burst characteristics more accurately capture pathophysiological and clinical features of the disease, with burst rate and inter-burst interval emerging as robust markers of disease state (29,70–72).

### No association between resting-state EEG parameters and motor disability

We further observed no significant associations between resting-state EEG parameters and either MDS-UPDRS-III scores or the amount of daily dopaminergic medication, indexed by LEDD. This finding is in line with previous studies reporting no relationship between aperiodic spectral parameters and motor symptom severity in either OFF or ON medication states (26,58), as well as evidence suggesting relatively modest effects of dopaminergic medication on resting-state spectral measures (although contrasting findings have also been reported (27)). Collectively, these observations suggest that the spectral alterations identified in the present study may reflect trait-like neurophysiological changes associated with PD rather than markers directly indexing momentary motor impairment or dopaminergic treatment burden.

Since neuroimaging studies demonstrated increased activity within sensorimotor cortical regions and altered functional interactions across cortical networks in patients with LID, even following withdrawal of dopaminergic medication (73,74), we explored whether previous exposure to dyskinesias might be associated with persistent alterations in cortical electrophysiological activity detectable in the OFF-medication state. Since we found no evidence for differences in resting-state spectral EEG parameters between patients with and without a history of LID, our findings do not support the presence of a latent resting-state spectral signature associated with a history of dyskinesias. Of note, EEG recordings were obtained exclusively in the OFF-medication state. Hence, they may not have captured medication-dependent alterations in cortico-basal ganglia network dynamics that have been demonstrated using pharmacodynamic neuroimaging approaches^21,61^.

### Age and disease duration effects on aperiodic activity

Although age-related changes in aperiodic spectral features have been described in healthy populations, their relationship with disease duration in PD remains largely unexplored (25). We found that the interaction between age and disease duration significantly predicted both aperiodic offset and exponent, whereas periodic alpha and beta power were unaffected. Specifically, longer disease duration was associated with higher offset and exponent values, with these effects becoming more pronounced at older ages. This observation is consistent with previous cross-sectional findings linking the aperiodic exponent to age in PD (58), and with recent longitudinal MEG evidence showing progressive increases in aperiodic offset and exponent that were associated with worsening motor symptoms and followed trajectories distinct from those observed in healthy controls(75). While age and disease duration are inherently related, the considerable variability in age at symptom onset suggests that they capture partially distinct processes. Our findings therefore support the notion that aperiodic spectral features may reflect the combined effects of ageing and PD-related neurodegeneration. Longitudinal studies using biologically defined disease staging and multimodal assessments are needed to determine whether these parameters represent reliable markers of disease progression (76).

### Methodological considerations

The cross-sectional design has inherent limitations. Prospective longitudinal EEG studies are needed to assess the temporal dynamics of altered resting-state cortical activity in PD, preferably starting at early prodromal stages of the disease. It also remains to be clarified how altered aperiodic resting-state activity in PD is modulated by motor or cognitive tasks and drugs boosting dopaminergic, noradrenergic, cholinergic or serotonergic neurotransmission.

Differences between PD and LID sub-groups investigated here are likely to be neglected by the OFF-medication state. More studies conducted recording neurophysiological data at the onset of dyskinesias or tracking dynamically the transition from the-OFF to the-ON state are warranted.

Spectral features were estimated using the FOOOF algorithm, which decomposes EEG power spectra into periodic and aperiodic components. While FOOOF toolbox offers the advantage of joint parameterization by simultaneously estimating both aperiodic and periodic components, alternative methods were not tested(77,78). Parameter choices were based on prior work and validated through visual inspection and goodness-of-fit metrics to minimize overfitting. We limited the number of peaks to two and set conservative thresholds to avoid fitting noise. Although cortical thickness reductions have been reported in non-demented PD(79), recent data indicate minimal influence of cortical thickness on spectral parameters estimates(29), suggesting that structural confounds are unlikely to explain our results.

## Conclusions

We identified robust resting-state EEG alterations in PD during the OFF-medication state, characterized by increased aperiodic offset and elevated alpha power relative to controls. Both offset and exponent, scaled with age and disease duration but not with UPDRS-III and LEDD, suggesting sensitivity to neurodegenerative progression beyond dopaminergic mechanisms. We conclude that spectral parameterization of EEG data offers a promising approach, able to feasibly and reliably characterize cortical dysfunction in PD, particularly in early or prodromal stages.

## Data Availability

All data produced in the present study are available upon reasonable request to the authors

## Acknowledgements

This study has been funded by the Lundbeck Foundation (Collaborative project grant “ADAptive and Precise Targeting of cortex-basal ganglia circuits in Parkinsońs Disease - ADAPT-PD”, grant nr. R336-2020-1035). The EEG data collection was supported by a research grant from The Danish Parkinson’s Association (Parkinsonforeningen). AGK was supported by research funds from the Danish Parkinson association (Grant No. A2479) and Capital Region of Denmark (Grant No. 2024-0050). DM was supported by a grant from the Danish Parkinson Association (Grant No. A1311), a research grant from Capital Region of Denmark (Grant No. A7656) and a Sapere Aude DFF-starting grant from the Independent Research Fund Denmark (Grant No. 1052-00054B). MCV was supported by The Danish Parkinson’s Association. BLT has been supported by Lundbeck Foundation, The Danish Parkinson’s Association and the Department of clinical medicine – University of Copenhagen.

## Authors Role

(1) Research Project: A. Design, B. Organization, C. Execution; (2) Statistical Analysis; A. Design, B. Execution, C. Review and Critique; (3) Manuscript Preparation: A. Writing of the first draft, B. Review and Critique.

S.B.: 1C, 2B-C, 3A

A.G.K.: 2A-B-C, 3B

D.M.: 2C, 3B

M.C.V.: 1C, 3B

N.R.: 1C, 3B

L.C.: 3C

B.L.T: 1C, 3B

A.L.: 3C

A.Q.: 3C

M.M.B.: 1A-B-C, 2A-C, 3B

H.R.S.: 1A, 2A-C, 3B

